# Genomic ascertainment of *CHEK2*-related cancer predisposition

**DOI:** 10.1101/2024.08.07.24311613

**Authors:** Sun Young Kim, Jung Kim, Mark Ramos, Jeremy Haley, Diane Smelser, H. Shanker Rao, Uyenlinh L. Mirshahi, Geisinger-Regeneron DiscovEHR Collaboration, Barry I. Graubard, Hormuzd A. Katki, David Carey, Douglas R. Stewart

## Abstract

**Purpose:** There is clear evidence that deleterious germline variants in *CHEK2* increases risk for breast and prostate cancers; there is limited or conflicting evidence for other cancers. Genomic ascertainment was used to quantify cancer risk in *CHEK2* germline pathogenic variant heterozygotes.

**Patients and Methods:** Germline *CHEK2* variants were extracted from two exome-sequenced biobanks linked to the electronic health record: UK Biobank (n= 469,765) and Geisinger MyCode (n=170,503). Variants were classified as per American College of Medical Genetics and Genomics (ACMG)/Association for Molecular Pathology (AMP) criteria. Heterozygotes harbored a *CHEK2* pathogenic/likely pathogenic (P/LP) variant; controls harbored benign/likely benign *CHEK2* variation or wildtype *CHEK2*. Tumor phenotype and demographic data were retrieved; to adjust for relatedness, association analysis was performed with SAIGE-GENE+ with Bonferroni correction.

**Results:** In *CHEK2* heterozygotes in both MyCode and UK Biobank, there was a significant excess risk of all cancers tested, including breast cancer (C50; OR=1.54 and 1.84, respectively), male genital organ cancer (C60-C63; OR=1.61 and 1.77 respectively), urinary tract cancer (C64-C68; OR=1.56 and 1.75, respectively) and lymphoid, hematopoietic, and related tissue cancer (C81-C96; OR=1.42 and 2.11, respectively). Compared to controls, age-dependent cancer penetrance in *CHEK2* heterozygotes was significantly younger in both cohorts; no significant difference was observed between the penetrance of truncating and missense variants for cancer in either cohort. Overall survival was significantly decreased in *CHEK2* heterozygotes in UK Biobank but there was no statistical difference in MyCode.

**Conclusion:** Using genomic ascertainment in two population-scale cohorts, this investigation quantified the prevalence, penetrance, cancer phenotype and survival in *CHEK2* heterozygotes. Tailored treatment options and surveillance strategies to manage those risks are warranted.

## Introduction

*CHEK2* (OMIM 604373) is a tumor-suppressor gene that encodes CHK2 (Serine/threonine protein kinase), involved in DNA repair in response to cellular DNA damage.^1^ There is clear evidence that deleterious germline variants in *CHEK2* heterozygotes are associated with an increased risk for female breast and prostate cancers; however, elevated risks for a variety of other cancers (e.g., colorectal, kidney, bladder, leukemia/lymphoma and thyroid) have been claimed but there is minimal, biased or conflicting evidence.^2^ In general, germline pathogenic truncating variants (PTV) (*e.g*., c.1100del p.(Thr367fs)) are associated with an increased risk of cancer. In contrast to PTV, pathogenic missense variants (PMV) in *CHEK2* have more variable effects, mainly dependent on whether a critical protein domain is affected. According to a study by Dorling et al.^3^, approximately 60% of rare PMV in *CHEK2* are associated with a lower risk of developing cancers compared to PTV. This suggests that the impact of PMV on cancer susceptibility is not uniform, but rather depends on the specific location and nature of the variants. To date, most work on quantifying risk from a germline variant in a cancer-predisposition gene has arisen from the well-established phenotype-first approach, in which individuals (and families) are ascertained from their presentation due to a clinical problem.

Genomic ascertainment is the inversion of the traditional phenotype-first approach^4^. With genomic ascertainment, germline variation of interest is identified, and phenotype status is then obtained from medical records to estimate variant prevalence and disease penetrance and characterize the phenotype. In principle, this should permit a less-biased estimate of the phenotypic spectrum, expressivity and penetrance of a deleterious variant or set of variants. Ascertainment biases still exist depending on how the cohort was recruited (healthier volunteer vs. clinical (health system or hospital) vs. true population sampling) that will influence risk estimates.

In this study, we used genomic ascertainment to quantify cancer risk for heterozygotes with germline pathogenic *CHEK2* variants. We analyzed electronic health record (EHR) in two population-based cohorts (UK Biobank (UKBB) and Geisinger MyCode) to estimate the prevalence, age-dependent penetrance, cancer risk and survival of *CHEK2* pathogenic heterozygotes compared to controls.

## Materials and Methods

### Cohorts and relatedness

From the UK Biobank, germline variants were obtained from field 23157, population level exome OQFE variants, and pVCF format, final exome release. Human subjects’ protection and review was through the North West Multi-centre Research Ethics Committee. Exome sequencing on UKBB samples has been described.^5,6^ The data was accessed January 2023; the number of unrelated participants was determined by R package ukbtools, “ukb_gene_samples_to_remove” function.

Geisinger is an integrated health system serving patients in Northeastern and Central Pennsylvania. All Geisinger patients are eligible to participate in the MyCode Community Health Initiative, a system-wide biorepository of blood and DNA samples for broad research purposes.^7^ Over 85% of Geisinger patients agree to participate and provided genomic data that are linked to their health records, which consist of routinely collected diagnosis, procedures, medication, and laboratory results, collected as part of their healthcare. This study was approved by the Geisinger Institutional Review Board. MyCode DNA samples were exome sequenced by the Regeneron Genetics Center using IDT exon capture probes as previously described.^8^ This study was approved by the Geisinger Institutional Review Board. In the Geisinger MyCode cohort, we included individuals over 18 years of age (n=167,050 with available exome data). To remove related individuals while maintaining the largest possible cohort, kinship pairs up to 3rd degree relatives (minimum PI_HAT = 0.1875) were used to create a graph of all relatives. Custom functions employing the network library in python were used. For each connected component (i.e., family), the node (i.e., patient) with the greatest number of edges (i.e., relatives) was removed. This was repeated until no edges remain in the connected component.

### Variant filtering and CHEK2 pathogenicity classification

Variants were filtered on the following quality metrics: Allelic Balance of Heterozygotes (ABHet) between 0.2 and 0.8, Genotype Quality >30, total read depth>5. All variants that pass quality metrics were annotated using snpEFF^9^, ANNOVAR^10^, ClinVar^11^ (database retrieved 09-23-2022), and InterVar (v.2.1.3)^12^. Variants were classified as pathogenic (P), likely pathogenic (LP), variant of uncertain significance (VUS), likely benign (LB), benign (B) using guidelines from the American College of Medical Genetics and Genomics and the Association for Molecular Pathology (ACMG/AMP).^13^ Final variant annotation was based on a hierarchical classification of ClinVar followed by InterVar^14^. “Heterozygotes” were defined as individuals who harbor a *CHEK2* P/LP variant, whereas controls included individuals who harbor canonical or B/LB *CHEK2* variation. *CHEK2* VUS were excluded. In this analysis, “All” refers to all *CHEK2* P/LP variants, “PTV” refers to predicted *CHEK2* truncating P/LP variants and “PMV” refers to pathogenic missense *CHEK2* P/LP variants. There were eight individuals and six individuals who harbored biallelic *CHEK2* variants in UKBB and MyCode, respectively; they were included in the All group, but were excluded from analyses of PTV and PMV. There was no individual who carried more than two P/LP variants in either UKBB or MyCode.

### Cancer phenotype and vital status query

Tumor phenotype and demographic data (age, sex, body mass index (BMI), alcohol consumption, smoking history, and race) were obtained for both heterozygotes and controls. Demographic comparisons were completed using Student T-test for continuous variables and Fisher’s exact test for binary variable. Clinical phenotypes of neoplasms were obtained using International Classification Diseases (ICD) diagnosis codes: ICD9 and ICD10 for UKBB, ICD10-Clinical Modification (CM) for MyCode data. The Geisinger Cancer Registry was also queried, which contains information on all patients diagnosed with cancer at a Geisinger facility; the Cancer Registry (field 40006 and 40013) and Death Registry data for UKBB (field 40001) were also utilized.

### Power to detect predisposition to common and rare cancers in UK Biobank and MyCode

Power estimates were performed by adapting formulas from Chow et al.^15^ to a cohort study setting with the assumption of non-biased ascertainment.

**Supplemental Figure 1** shows power as a function of presumed true odds ratio for a range of cancer rates in the UK Biobank and MyCode cohorts using cohort-specific All, PTV and PMV *CHEK2* heterozygote prevalence from **Table 1**. For All, PTV and PMV *CHEK2* heterozygotes, there is 100% power in both UK Biobank and MyCode to detect common cancers (≥5% cancer rate, which include many sex-specific cancers such as female breast and prostate) with an odds ratio of >2. For All, PTV and PMV *CHEK2* heterozygotes, there is ≥80% power to detect rare cancers (≥1% cancer rate) with an odds ratio of >2. For All *CHEK2* heterozygotes, there is >80% power to detect very rare cancers (≥0.1%) with an odds ratio of >2.7 in both MyCode and UKBB; there is less power in the PTV- and PMV-specific cohorts.

**Table 1.** Prevalence of All, pathogenic truncating variants (PTV) and pathogenic missense variants (PMV) *CHEK2* in adult heterozygotes in UK Biobank and Geisinger MyCode. The sum of PTV and PMV is less than All total due to presence of non-canonical splice-site variants.

| Cohort | Individuals/Prevalence<br>(95%CI) | All *CHEK2* P/LP Variants | Pathogenic Truncating<br>Variants (PTV) | Pathogenic Missense<br>Variants (PMV) |
| --- | --- | --- | --- | --- |
| **UK Biobank – related**<br>**and unrelated**<br>**(n=469,765)** | Number of individuals | 3,232 | 1,847 | 1,290 |
|  | Prevalence | 1/145 (1/140 – 1/150) | 1/254 (1/243 – 1/266) | 1/364 (1/344 – 1/384) |
| **UK Biobank –**<br>**unrelated (n=437,645)** | Number of individuals | 3,171 | 1,825 | 1,268 |
|  | Prevalence | 1/138 (1/133-1/142) | 1/239 (1/229-1/251) | 1/345 (1/326-1/364) |
| **MyCode – related and**<br>**unrelated (n=167,050)** | Number of individuals | 3,153 | 913 | 2,221 |
|  | Prevalence | 1/52 (1/51 – 1/54) | 1/183 (1/171 – 1/195) | 1/75 (1/72 – 1/78) |
| **MyCode – unrelated**<br>**(n=109,730)** | Number of individuals | 2,489 | 728 | 1,751 |
|  | Prevalence | 1/43 (1/41 – 1/44) | 1/150 (1/140-1/162) | 1/62 (1/59-1/65) |

### Cancer risk estimate

Cancer prevalence was modeled using logistic regression with carrier status for All, PTV, and PMV as the main set of explanatory variables and age, sex, smoking history, alcohol consumption and BMI as covariates. For sex-specific cancers (C51-C58 for female; C60-C63 for male) prevalence was analyzed only with female or male controls. Multiplicity issues were addressed using Bonferroni adjustment at family-wise error rate of α=0.05. To correct for relatedness, we used SAIGE-GENE+ version 1.1.6.2. Covariates include using PC1-4, current age, sex, smoking, alcohol use and BMI.^16^ To further help guard against inaccurate p-values and confidence interval coverage for associations arising from very low prevalence of rare cancers, cancers where there were less than five cases among heterozygotes were excluded from any analyses.

### Kaplan-Meier, cancer penetrance and mortality in individuals with cancer

Kaplan-Meier survival analyses were used to estimate all-cause mortality, penetrance of pathogenic *CHEK2* variants for cancer, and overall survival for individuals with cancer in MyCode and UKBB cohorts. In the MyCode cohort, only events occurring 3 months or after in Geisinger facilities are included in survival analyses. Data was truncated to individuals with current age ≤ 85. Hazard ratios were computed using Cox Proportional-Hazards (coxph) model adjusting for age, self-reported race, sex, smoking history, alcohol consumption and BMI, using Log-rank test for equality to compare differences between the curves for controls and variant groups. Coxph also adjusted for relatedness by clustering genetically inferred family units. All the analyses were conducted using R version 4.1.2.

## Results

### Prevalence and demographics of All, PTV and PMV CHEK2 heterozygotes in MyCode and UK Biobank

**Table 1** shows the prevalence of All, PTV and PMV *CHEK2* heterozygotes in both cohorts. (The sum of PTV and PMV is less than All total due to the presence of non-canonical splice-site variants; **Supplemental Table 1** provides details on the variants) The relatedness (up to the third degree) of the MyCode and UKBB cohorts is ∼30% and ∼10% respectively; **Table 1** also shows the heterozygote prevalence in the unrelated fraction of the two cohorts. **Supplemental Table 2** lists demographics and covariates between All, PTV and PMV heterozygotes and controls. There were 305,330 controls (65%) in UKBB and 152,662 controls (91%) in MyCode.

### Significant excess risk for cancers of the breast, male genital organ, urinary tract and lymphoid, hematopoietic, and related tissues in CHEK2 heterozygotes in both MyCode and UKBB

Figure 1A displays statistically significant association of pathogenic *CHEK2* All, PTV and PMV for organ system groupings of cancer in MyCode. The odds ratios and Bonferroni-corrected p-values for the association between *CHEK2* heterozygotes for organ system groupings of cancer ICD codes are shown. In All *CHEK2* heterozygotes, there was a significant excess risk (Bonferroni-adjusted SAIGE p-value) of all cancers, breast cancer (C50), male genital organ cancer (C60-C63), urinary tract cancer (C64-C68), thyroid and other endocrine gland cancer (C73-C75), and lymphoid, hematopoietic, and related tissue cancer (C81-C96). (Of all the C50 codes observed in *CHEK2* heterozygotes, 155 and 225 (99.4, 99.1%) were in females and, 1 and 2 (0.6, 0.8%) were in males in MyCode and UK Biobank, respectively.) **Supplemental** Figure 2 displays the odds ratio for the MyCode cohort for All, PTV and PMV *CHEK2* heterozygotes for all organ system groupings of cancer ICD codes. Figure 1B displays the odds ratio for the UKBB cohort for All, PTV and PMV *CHEK2* heterozygotes for organ system groupings of cancer ICD codes with a significant excess of risk. In All *CHEK2* heterozygotes, there was a significant excess risk (Bonferroni-adjusted SAIGE p-value) of developing all cancers, breast cancer (C50), male genital organ cancer (C60-C63), urinary tract cancer (C64-C68), cancer from the secondary and unspecified sites (C76-C79), and lymphoid, hematopoietic, and related tissue cancer (C81-C96). In contrast to MyCode, there was a non-significant excess risk to develop thyroid and other endocrine gland cancer (C73-C75). **Supplemental** Figure 3 displays the odds ratio for the UKBB cohort for All, PTV and PMV *CHEK2* heterozygotes for all organ system groupings of cancer ICD codes.

**Figure 1.**
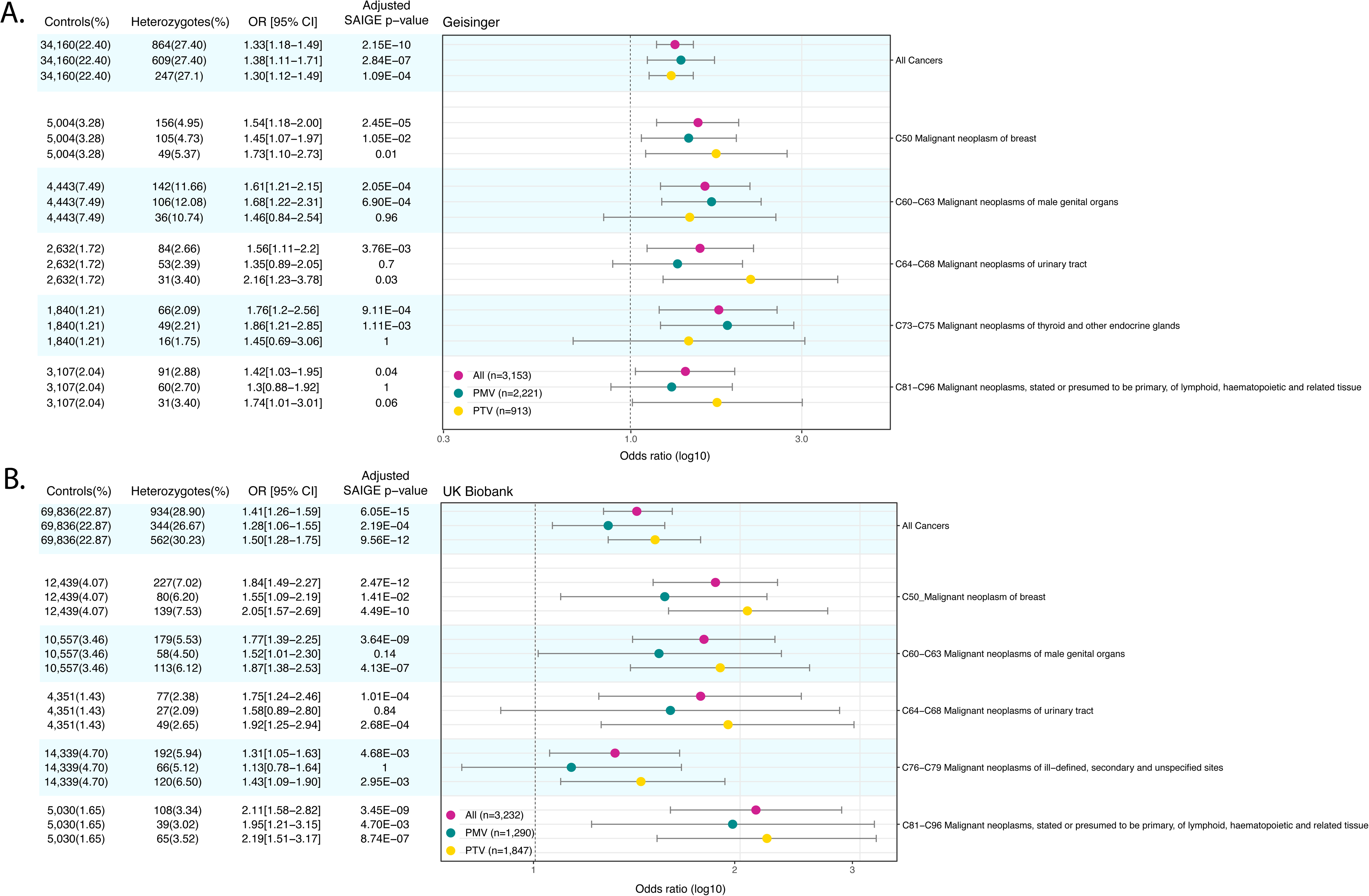
Odds ratio for All, PTV and PMV *CHEK2* heterozygotes for organ system groupings of cancer ICD codes with a significant excess of risk in MyCode (panel A) and UK Biobank (panel B). CI: 95% confidence interval; OR: odds ratio; PMV: pathogenic missense variant; PTV: pathogenic truncating variant

### Specific cancers related to All, PTV and PMV CHEK2 heterozygotes

Figure 2A shows the specific types of cancer in the MyCode cohort with an excess risk from the significant organ-system analysis shown in Figure 1A. Of note is the significant excess risk for prostate cancer (C61), kidney cancer (C64), bladder cancer (C67), thyroid cancer (C73), and lymphoid leukemia (C91) in All heterozygotes. **Supplemental** Figure 4 displays the odds ratio for the MyCode cohort for All, PTV and PMV *CHEK2* heterozygotes for all specific types of cancer from all organ system groupings of cancer ICD codes. **Supplemental Table 3** lists the case counts and percentages for PMV, PTV and All cohorts and fold-enrichment (vs. controls) for each of the ICD10 diagnostic codes in MyCode.

**Figure 2.**
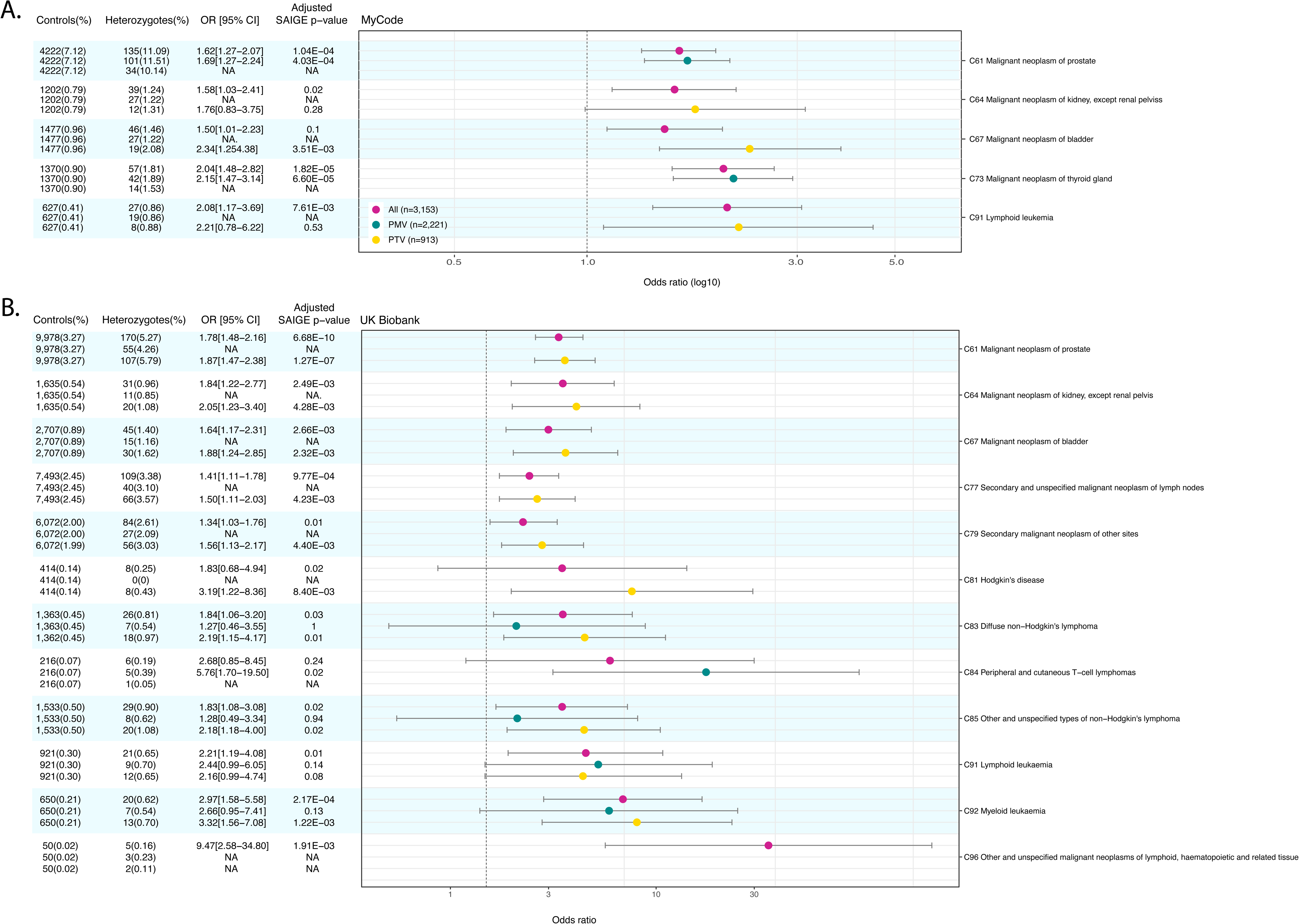
Odds ratio for All, PTV and PMV *CHEK2* heterozygotes for specific cancers in the organ system groupings of cancer ICD codes with a significant excess of risk in MyCode (panel A) and UK Biobank (panel B). CI: 95% confidence interval; OR: odds ratio; PMV: pathogenic missense variant; PTV: pathogenic truncating variant

Figure 2B shows the specific types of cancer in the UKBB cohort with an excess risk from the organ-system analysis shown in Figure 1B. Of note is the significant excess risk for prostate (C61), kidney cancer (C64), and bladder cancer (C67) in All and PTV heterozygotes. There was significant increased risk for diffuse non-Hodgkin lymphoma (C83), other and non-specified types of non-Hodgkin lymphoma (C85) and lymphoid leukemia (C91) in All heterozygotes, whereas peripheral and cutaneous T-cell lymphomas (C84) were exclusively associated with PMV heterozygotes. **Supplemental** Figure 5 displays the odds ratio for the UK Biobank cohort for All, PTV and PMV *CHEK2* heterozygotes for all specific types of cancer from all organ system groupings of cancer ICD codes. **Supplemental Table 3** lists the case counts and percentages for PMV, PTV and All cohorts and fold-enrichment (vs. controls) for each of the ICD10 diagnostic codes in UK Biobank.

### Age-dependent penetrance differs significantly in All, PTV and PMV CHEK2 heterozygotes vs. controls, but not between CHEK2 PTV vs. PMV heterozygotes

Compared to controls, age-dependent penetrance in All *CHEK2* variants for all cancers was significantly different in both MyCode (adjusted HR: 1.26 [95%CI 1.17-1.36], P-value: 6.1x10^-10^) and UKBB (adjusted HR 1.31 [95%CI 1.24-1.40], *P*-value: 2.0x10^-16^) (Figures 3A and **4A**). *CHEK2* PMV or PTV heterozygotes alone were at higher risk for all cancers tested compared to controls in both MyCode and UKBB. In MyCode, for PMV (vs. controls) the adjusted HR: 1.24 [1.13-1.35], p value=2.71x10^-06^; in UKBB, for PMV (vs. controls) the adjusted HR: 1.17 [1.06-1.30], p-value=1.56x10^-3^. In MyCode, for PTV (vs. controls) adjusted HR: 1.30 [1.13-1.50], p value=2.1x10^-4^; in UKBB for PTV (vs. controls) the adjusted HR:1.34 [1.23-1.45], p-value=1.67x10^-12^. There was no significant difference in the penetrance of *CHEK2* PTV vs. PMV for cancers in MyCode (univariate HR: 1.06 [0.90-1.25], *P*-value=0.47) and the UKBB (adjusted HR 1.15 [95%CI 1.00-1.33], *P*-value: 0.05).

**Figure 3.**
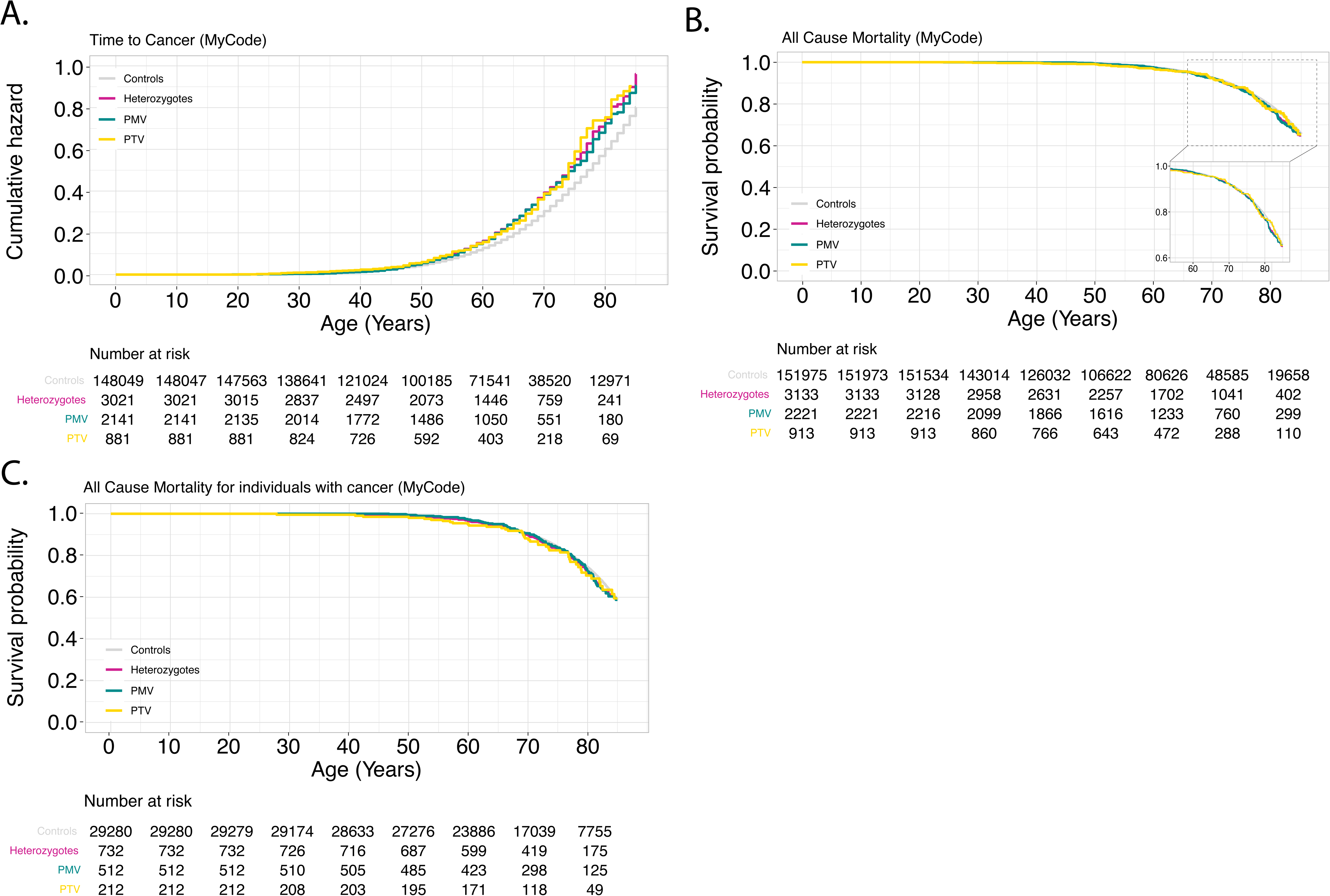
Penetrance of pathogenic *CHEK2* variants for cancer and all-cause mortality in MyCode. **Panel A**: Time-to-cancer (penetrance); **Panel B**: All-cause mortality; **Panel C**: All-cause mortality for individuals with cancer. PMV: pathogenic missense variant; PTV: pathogenic truncating variant.

**Figure 4.**
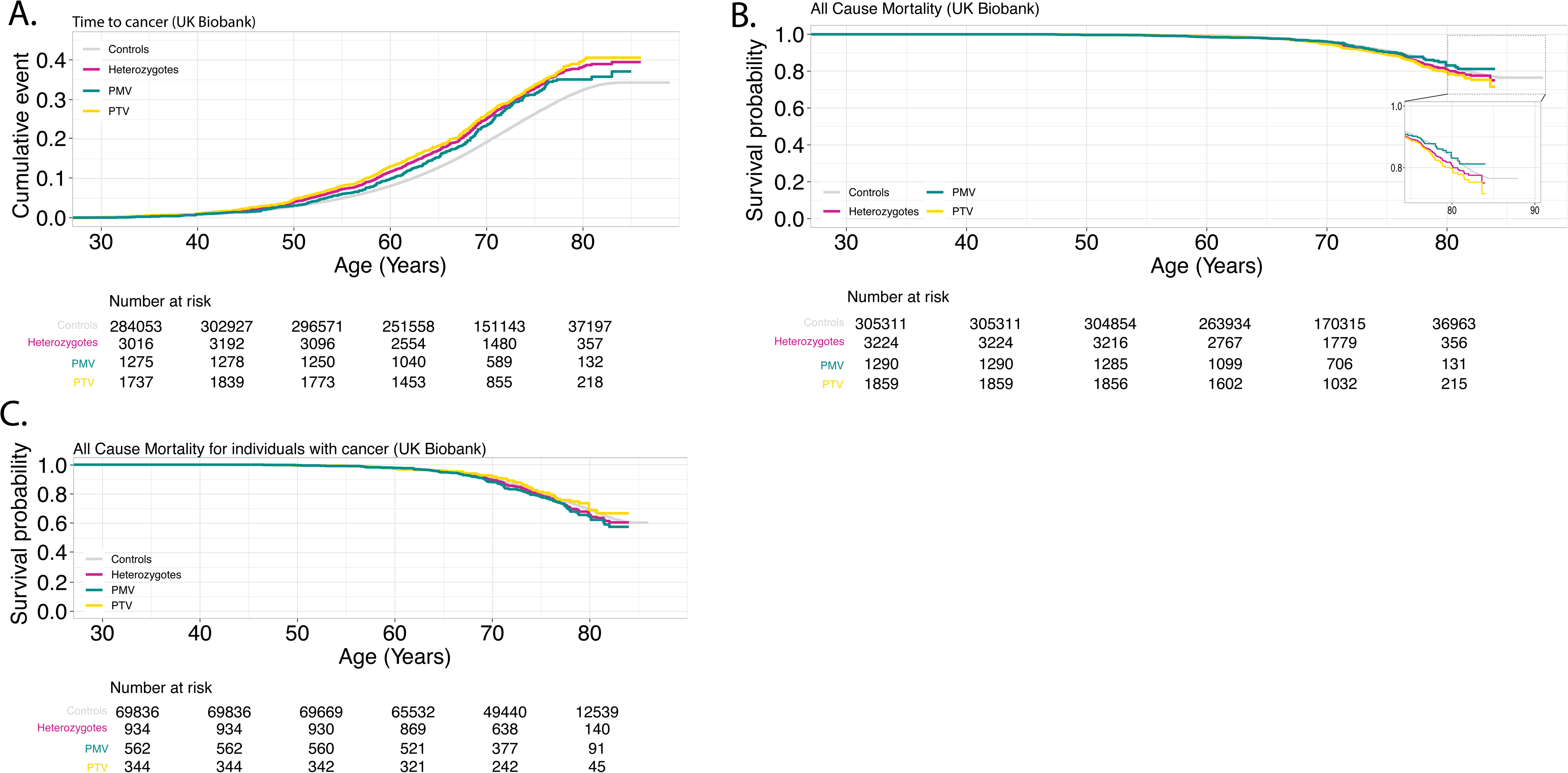
Penetrance of pathogenic *CHEK2* variants for cancer and all-cause mortality in UK Biobank. **Panel A**: Time-to-cancer (penetrance); **Panel B**: All-cause mortality; **Panel C**: All-cause mortality for individuals with cancer. PMV: pathogenic missense variant; PTV: pathogenic truncating variant

### All-cause mortality was significantly increased in All heterozygotes compared to controls in UKBB but not MyCode

All-cause mortality was significantly increased in All heterozygotes in UKBB (adjusted HR 1.21 [95%CI 1.08-1.37], P-value: 1.51E-3) but not in MyCode (adjusted HR 1.09 [95%CI 0.96-1.24], *P*-value: 0.20) (Figures 3B and **4B**). There was no significant difference in all-cause mortality in PTV and PMV heterozygotes in UKBB (adjusted HR 1.24 [95%CI 0.97-1.60], *P*-value: 0.10) and MyCode (adjusted HR 1.06 [95%CI 0.80-1.41], *P*-value: 0.67).

### All-cause mortality amongst individuals with cancer was not significantly increased in All heterozygotes compared to controls in UKBB and MyCode

There was no statistical difference between All heterozygotes and controls in both MyCode (adjusted HR 1.08 [95%CI 0.90-1.30], *P*-value=0.43) and the UKBB (adjusted HR 1.12 [95%CI 0.98-1.29], *P*-value: 0.11) cohorts for all-cause mortality in individuals with cancer. There were no significant differences between PTV and PMV heterozygotes in either the MyCode (adjusted HR 1.20 [95%CI 0.81-1.78], *P*- value=0.35) or UKBB (adjusted HR 1.33 [95%CI 0.98-1.80], *P*-value: 0.07) cohorts (Figure 3C and **4C**).

## Discussion

In this investigation, familial relationship-adjusted, Bonferroni-corrected genomic ascertainment of two population-based, exome-sequenced, EHR-linked cohorts was used to quantify risk of cancers arising from pathogenic/likely pathogenic germline variants in *CHEK2*. Notably, given the stated assumptions about participant ascertainment, both cohorts had high power to detect elevated risk (OR>2) in all but the rarest cancers. Genomic ascertainment quantifies risk based on genotype (not phenotype) and thus may reduce risk inflation arising from cancer ascertainment (case/family recruitment) by personal and/or family medical history.

Clinically, this investigation confirms the significantly increased risk for breast and prostate cancers (as well as all cancers, collectively), although the observed risk tends to be even lower (OR<2) than previous estimates, especially for PTV (typically OR>2).^2^ Interestingly, in neither cohort was a significant excess risk for “malignant neoplasms of digestive organs” (majority were colorectal cancers) observed for All, PTV or PMV (**Supplemental Table 3**). Published risk estimates for colorectal cancer from *CHEK2* PTV are more modest (OR ∼2) and conflicting than those for female breast cancer and prostate cancer; higher estimates of risk are driven by studies of multiplex families.^17^ Published risk for colorectal cancer from *CHEK2* PMV tend to be even lower (OR<2) or non-significant.^2,18^ Given this, a recent ACMG review and clinical practice guideline on management^2^ concluded that *CHEK2* heterozygosity is not clinically actionable for colorectal cancer risk in isolation and to offer surveillance as per family history. In contrast, current National Comprehensive Cancer Network (NCCN) guidelines recommend colorectal cancer screening for individuals who carry *CHEK2* P/LP variants. (https://www.nccn.org/professionals/physician_gls/pdf/genetics_colon.pdf) Our observations in this study of non-significant colorectal risk are congruent with the ACMG recommendations. In summary, although additional confirmation is needed for breast, prostate and colorectal cancers, genomic ascertainment showed generally lower (or non-significant) risk than previously reported for All, PTV and PMV in *CHEK2*.

This work provides substantial evidence from both cohorts of significant increased risk for kidney cancer, bladder cancer and CLL (lymphoid leukemia). In this investigation, Bonferroni correction was applied to organ-system groupings and not specific cancer types. Thus, other cancers may be enriched in *CHEK2* heterozygotes; **Supplemental Table 2** lists counts of cancer types in controls and All, PTV and PMV heterozygotes. Recent ACMG clinical practice guideline on management of *CHEK2* heterozygotes^2^ concluded that there was likely an increased risk for kidney cancer but that larger studies with appropriate controls were needed. Several publications found a range of risk (OR=3; hazard ratio =10.8)^18–21^; other investigations had non-significant findings.^22^ As with breast and prostate cancers in this study, genomic ascertainment resulted in lower risk estimates (OR<2) for kidney cancer than previous studies and was remarkably consistent across the two cohorts. A 2023 ACMG review and clinical guidance for *CHEK2* heterozygotes^2^ noted a single publication of non-significant *CHEK2*-associated bladder cancer^23^ but deemed this evidence insufficient to make recommendations; more recent publications have found additional evidence of a *CHEK2*-bladder cancer association.^24,25^ Genomic ascertainment in this study revealed similarly increased bladder cancer risk in both cohorts (especially in PTV). In summary, this evidence of increased risk for both renal and bladder cancers should prompt the development of clinical management recommendations for surveillance and intervention for these cancers.

In both cohorts there was significantly elevated risk for lymphoid and hematopoietic neoplasms collectively (C81-C96); across all the subtypes of these malignancies, only CLL (lymphoid leukemia) had significantly elevated risk (>2) in both cohorts. Reports of increased risk of hematologic malignancy (especially CLL) in *CHEK2* heterozygotes date from 2006^26,27^ but were conflicting and/or based on highly ascertained families. A 2022 investigation using a PheWAS approach in an earlier version of UKBB reported an excess risk (OR>3) for leukemia and plasma cell neoplasms in *CHEK2* P/LP heterozygotes.^28^ To date with current approaches (*e.g.,* CBC, physical exam) there is limited evidence-based actionability for surveillance for increased risk of leukemia, however with developing methods (*e.g.,* methylation profiling of circulating tumor DNA) this may improve. Outcomes and tailored treatment options for *CHEK2*-associated CLL merit investigation.

A significant excess of malignancies of thyroid and other endocrine tumors (C73-C75) was observed in MyCode but not UK Biobank; this was almost entirely driven by thyroid tumors (C73) and, unlike most other associations, by *CHEK2* PMV. Previous studies have been conflicting or limited by small numbers or single-country ascertainment.^18,22,29^ The recent ACMG review and clinical guidance for *CHEK2* heterozygotes^2^ did not find sufficient evidence to support a clear association for thyroid cancer and did not recommend surveillance. Genomic ascertainment of *DICER1*-associated thyroid disease (*e.g.,* goiter) also found significant differences in *DICER1* heterozygotes (vs. controls) in MyCode but not UK Biobank and may reflect the different medical cultures in the US and UK in approaches to medical imaging of the thyroid.^30^ Conversely, there was a significant excess risk of “malignant neoplasms of ill-defined, secondary and unspecified sites” (C76-C79) in UK Biobank but not MyCode.

Numerous other associations have been observed for specific cancers for *CHEK2* heterozygotes including sarcoma, stomach, male breast, melanoma, endometrial and testicular cancer^2^. For more common cancers (endometrial, skin), there was no evidence of association for these in either cohort. For some rarer cancers (male breast, testicular) the two cohorts were likely underpowered (**Supplemental** Figure 1); for others (sarcoma, stomach) there may be both a power issue and a survival bias in ascertainment given the aggressive nature of these cancers.

Overall, pathogenic germline *CHEK2* All, PTV and PMV are common, but the conferred excess cancer risk is, with few exceptions, less than an OR of 2. In addition, the lack of significant difference between *CHEK2* All heterozygotes and controls in all-cause mortality in individuals with cancer suggests that germline *CHEK2*-associated cancer is not clinically more aggressive than non-*CHEK2*-associated cancer. The degree of risk from PTV and PMV overlap considerably with risk of PMV generally lower. The clinical relevance of this may be debatable since penetrance for cancer, all-cause mortality and all-cause mortality in individuals with cancer was not significantly different between PMV and PTV in both cohorts.

There are limitations to these retrospective analyses. MyCode and UK Biobank are predominantly of European ancestry. Copy-number (deletions) in *CHEK2* were not evaluated due to limited data availability in UK Biobank. Enrollment in the two cohorts was subject to ascertainment biases as individuals with conditions leading to death or disabilities would be less likely to participate. The “healthy volunteer” bias (compared to the UK population) of the UK Biobank has been documented ^31^

In summary, we quantified cancer risk and survival in *CHEK2* heterozygotes using the novel genome-first approach in two well-powered cohorts. Our findings inform clinical care by supporting current recommendations for prostate and breast cancer surveillance and provide definitive evidence of increased risk for renal, bladder, and CLL in heterozygotes with pathogenic *CHEK2* variants. Tailored treatment options and surveillance strategies to manage those risks are needed.

The content of this publication does not necessarily reflect the views or policies of the Department of Health and Human Services, nor does mention of trade names, commercial products or organizations imply endorsement by the U.S. Government.

The authors would like to acknowledge the participants of the MyCode Community Initiative for the use of their health and genomic information, without whom this study would not be possible. The patient enrollment and exome sequencing were funded by the Regeneron Genetics Center. Data for this project was made possible by the Geisinger-Regeneron DiscovEHR Collaboration.

## Data Availability

All data produced are available online at https://www.ukbiobank.ac.uk/

## Acknowledgments

This work was supported by the Intramural Research Program of the Division of Cancer Epidemiology and Genetics of the National Cancer Institute, Bethesda, MD and utilized the computational resources of the NIH High-Performance Computing Biowulf cluster. This research has been conducted using the UK Biobank Resources under application 54389.

## Supplemental Figures

**Supplemental Figure 1.** Power as a function of risk (odds ratio) in MyCode (**Panel A, C, E**) and UK Biobank (**Panels B, D, F**) for a range of cancer rates. Prevalence data from cohort-specific ALL (Panels A, B) pathogenic truncating variants (PTV) (Panels C, D) and pathogenic missense variants (PMV) (Panels E, F) *CHEK2* heterozygotes (Table 1). Dark gray line represents 80% power, and light gray line represents 90% power.

**Supplemental Figure 2.** Odds ratio for All, PTV and PMV *CHEK2* heterozygotes for organ system groupings of cancer ICD codes in MyCode. Red font represents significant cancers. CI: 95^%^ confidence interval; OR: odds ratio; PMV: pathogenic missense variant; PTV: pathogenic truncating variant

**Supplemental Figure 3.** Odds ratio for All, PTV and PMV *CHEK2* heterozygotes for organ system groupings of cancer ICD codes in UK Biobank. Red font represents significant cancers. CI: 95^%^ confidence interval; OR: odds ratio; PMV: pathogenic missense variant; PTV: pathogenic truncating variant

**Supplemental Figure 4.** Odds ratio for All, PTV and PMV *CHEK2* heterozygotes for all specific cancers in the organ system groupings of cancer ICD codes in MyCode. Red font represents significant cancers. CI: 95^%^ confidence interval; OR: odds ratio; PMV: pathogenic missense variant; PTV: pathogenic truncating variant

**Supplemental Figure 5.** Odds ratio for All, PTV and PMV *CHEK2* heterozygotes for all specific cancers in the organ system groupings of cancer ICD codes in UK Biobank. Red font represents significant cancers. CI: 95^%^ confidence interval; OR: odds ratio; PMV: pathogenic missense variant; PTV: pathogenic truncating variant

## Supplemental Tables

**Supplemental Table 1. List of all variants found in the study**

**Supplemental Table 2. Demographics of *CHEK2* heterozygotes vs. controls**

**Supplemental Table 3. Case counts and percentages for PMV, PTV and All cohorts and fold-enrichment (vs. controls) for each of the ICD10 diagnostic codes in MyCode and UK Biobank.**

