## Supplementary figures and images for "Genomic ascertainment of *CHEK2*-related cancer predisposition"

### Supplemental Figure 1

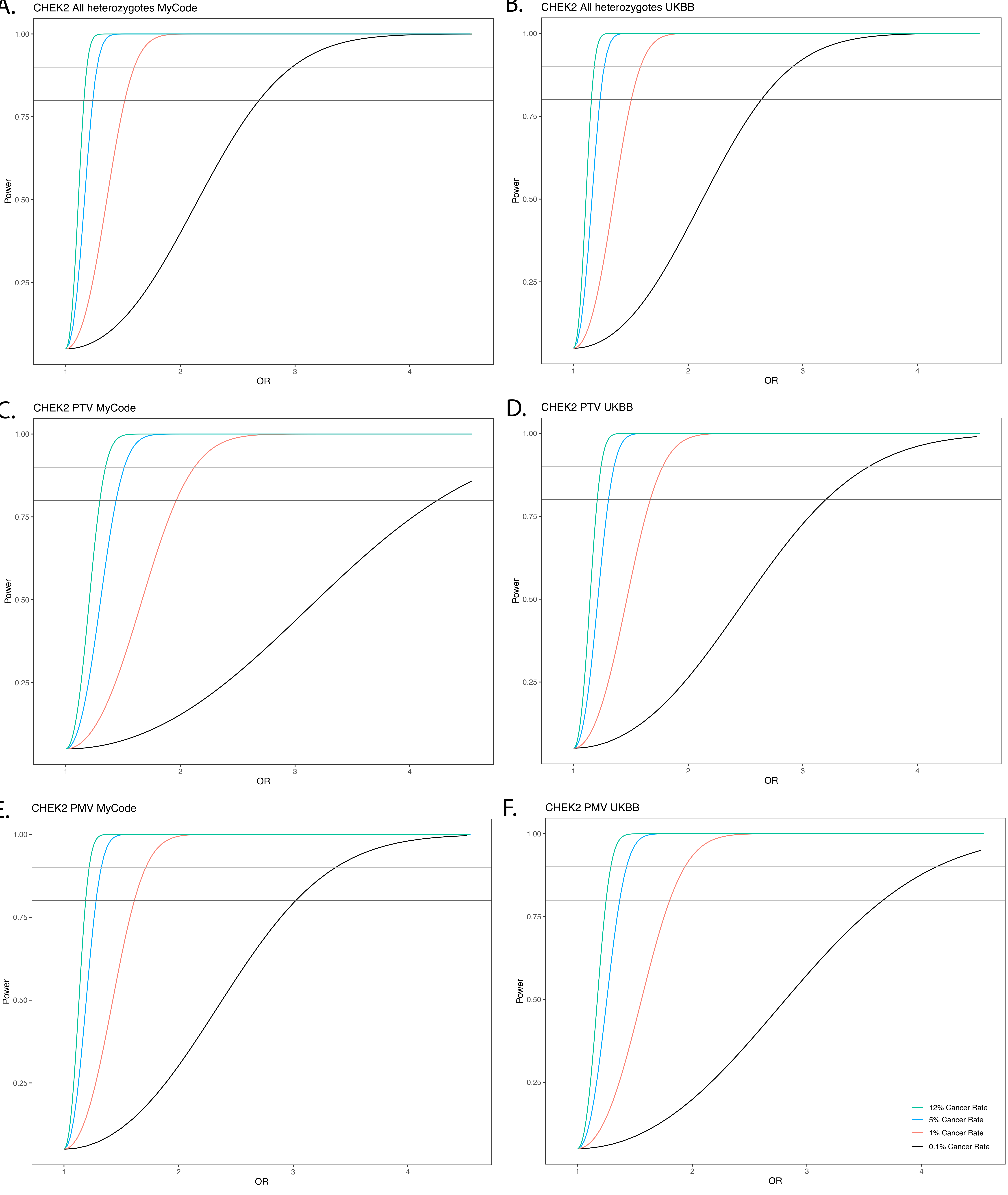

### Supplemental Figure 2

| Controls(%) | Heterozygotes(%) | OR [95% CI] | Adjusted SAIGE p-value |
|---|---|---|---|
| 34,160(22.40) | 864(27.40) | 1.33[1.18–1.49] | 2.15E–10 |
| 34,160(22.40) | 609(27.40) | 1.38[1.11–1.71] | 2.84E–07 |
| 34,160(22.40) | 247(27.1) | 1.30[1.12–1.49] | 1.09E–04 |
| 895(0.59) | 21(0.67) | 1.1[0.56–2.13] | 1 |
| 895(0.59) | 12(0.54) | 0.87[0.37–2.03] | 1 |
| 895(0.59) | 9(0.99) | 1.71[0.63–4.65] | 1 |
| 4,096(2.68) | 109(3.48) | 1.27[0.94–1.7] | 0.33 |
| 4,096(2.68) | 78(3.51) | 1.26[0.89–1.78] | 0.14 |
| 4,096(2.68) | 30(3.29) | 1.26[0.72–2.21] | 1 |
| 2,791(1.83) | 54(1.71) | 0.9[0.6–1.37] | 1 |
| 2,791(1.83) | 38(1.71) | 0.88[0.53–1.45] | 1 |
| 2,791(1.83) | 16(1.75) | 0.97[0.45–2.1] | 1 |
| 256(0.17) | 7(0.22) | 1.31[0.42–4.09] | 1 |
| 256(0.17) | 6(0.27) | 1.57[0.7–3.54] | 1 |
| 256(0.17) | 1(0.11) | NA | NA |
| 12,443(8.15) | 252(7.99) | 0.95[0.77–1.17] | 1 |
| 12,443(8.15) | 174(7.83) | 0.9[0.71–1.15] | 1 |
| 12,443(8.15) | 76(8.32) | 1.05[0.72–1.53] | 1 |
| 624(0.41) | 15(0.47) | 1.15[0.53–2.49] | 1 |
| 624(0.41) | 13(0.59) | 1.39[0.8–2.42] | 1 |
| 624(0.41) | 1(0.11) | NA | NA |
| 5,004(3.28) | 156(4.95) | 1.54[1.18–2.00] | 2.45E–05 |
| 5,004(3.28) | 105(4.73) | 1.45[1.07–1.97] | 1.05E–02 |
| 5,004(3.28) | 49(5.37) | 1.73[1.10–2.73] | 0.01 |
| 3,016(3.23) | 66(3.41) | 1.05[0.72–1.54] | 1 |
| 3,016(3.23) | 45(3.35) | 1.01[0.64–1.59] | 1 |
| 3,016(3.23) | 20(3.46) | 1.11[0.56–2.19] | 1 |
| 4,443(7.49) | 142(11.66) | 1.61[1.21–2.15] | 2.05E–04 |
| 4,443(7.49) | 106(12.08) | 1.68[1.22–2.31] | 6.90E–04 |
| 4,443(7.49) | 36(10.74) | 1.46[0.84–2.54] | 0.96 |
| 2,632(1.72) | 84(2.66) | 1.56[1.11–2.2] | 3.76E–03 |
| 2,632(1.72) | 53(2.39) | 1.35[0.89–2.05] | 0.7 |
| 2,632(1.72) | 31(3.40) | 2.16[1.23–3.78] | 0.03 |
| 1,353(0.89) | 42(1.33) | 1.49[0.93–2.38] | 0.42 |
| 1,353(0.89) | 33(1.49) | 1.65[0.99–2.76] | 0.14 |
| 1,353(0.89) | 8(0.88) | 0.99[0.34–2.86] | 1 |
| 1,840(1.21) | 66(2.09) | 1.76[1.2–2.56] | 9.11E–04 |
| 1,840(1.21) | 49(2.21) | 1.86[1.21–2.85] | 1.11E–03 |
| 1,840(1.21) | 16(1.75) | 1.45[0.69–3.06] | 1 |
| 5,693(3.73) | 152(4.82) | 1.28[0.99–1.66] | 0.09 |
| 5,693(3.73) | 104(4.68) | 1.22[0.9–1.66] | 0.84 |
| 5,693(3.73) | 46(5.04) | 1.4[0.89–2.2] | 0.64 |
| 3,107(2.04) | 91(2.88) | 1.42[1.03–1.95] | 0.04 |
| 3,107(2.04) | 60(2.70) | 1.3[0.88–1.92] | 1 |
| 3,107(2.04) | 31(3.40) | 1.74[1.01–3.01] | 0.06 |

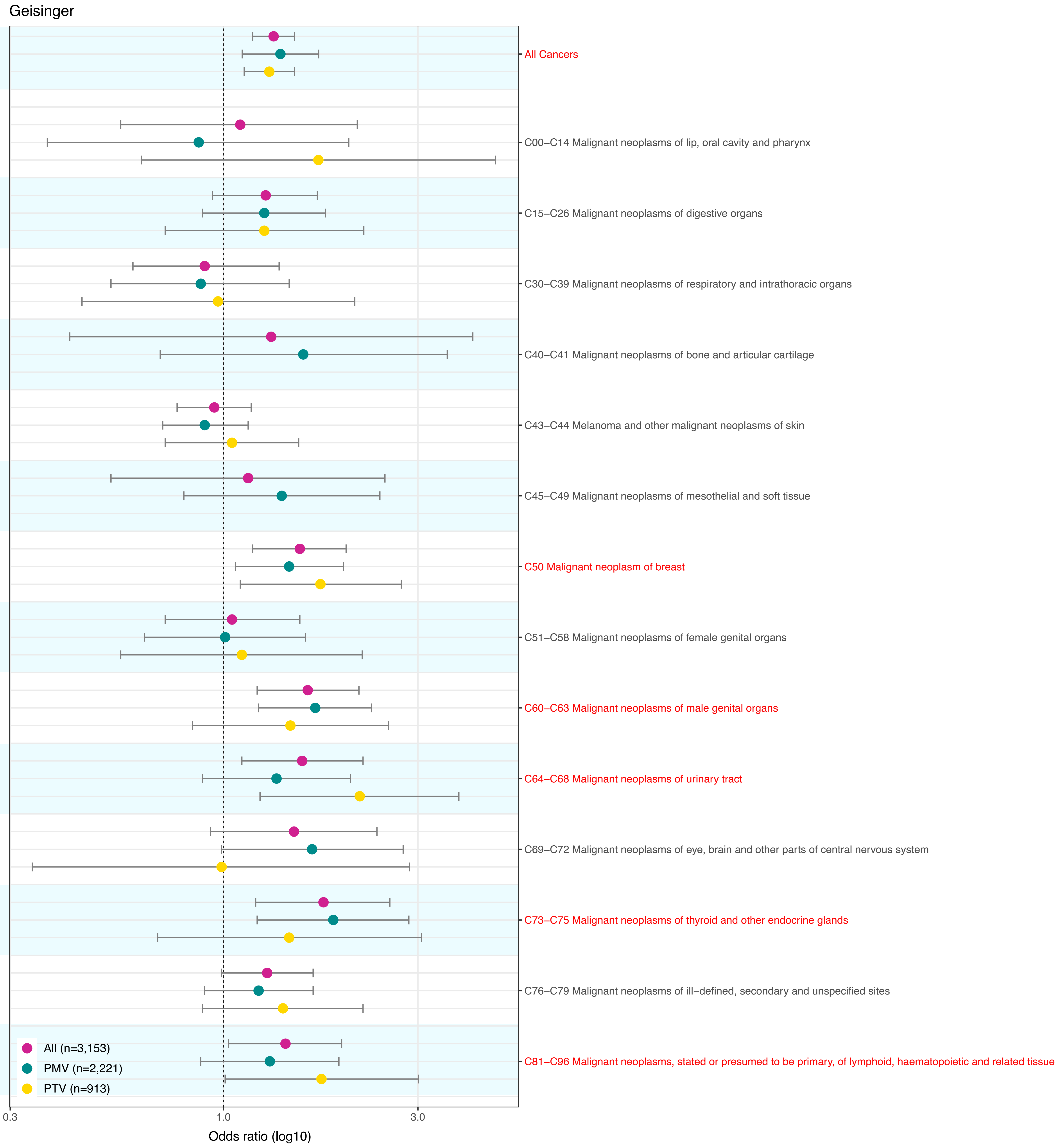
