## Supplemental Figure 3 for "Genomic ascertainment of *CHEK2*-related cancer predisposition"

| Controls(%) | Heterozygotes(%) | OR [95% CI] | Adjusted SAIGE p−value | |
|---|---|---|---|---|
| 69,836(22.87) | 934(28.90) | 1.41[1.26−1.59] | 6.05E−15 | |
| 69,836(22.87) | 344(26.67) | 1.28[1.06−1.55] | 2.19E−04 | All Cancers |
| 69,836(22.87) | 562(30.23) | 1.50[1.28−1.75] | 9.56E−12 | |
| 1,270(0.42) | 10(0.31) | 0.75[0.30−1.88] | 1 | |
| 1,270(0.42) | 4(0.31) | NA | NA | C00−C14 Malignant neoplasms of lip, oral cavity and pharynx |
| 1,270(0.42) | 6(0.32) | 0.78[0.24−2.57] | 1 | |
| 10,048(3.29) | 115(3.55) | 1.11[0.84−1.47] | 1 | |
| 10,048(3.29) | 44(3.41) | 1.09[0.69−1.71] | 1 | C15−C26 Malignant neoplasms of digestive organs |
| 10,048(3.29) | 66(3.57) | 1.09[0.76−1.58] | 1 | |
| 4,169(1.37) | 44(1.36) | 1.03[0.66−1.61] | 1 | |
| 4,169(1.37) | 18(1.40) | 1.05[0.52−2.11] | 1 | C30−C39 Malignant neoplasms of respiratory and intrathoracic organs |
| 4,169(1.37) | 24(1.30) | 0.98[0.54−1.79] | 1 | |
| 218(0.07) | 1(0.03) | NA | NA | |
| 218(0.07) | 0(0) | NA | NA | C40−C41 Malignant_neoplasms of bone and articular cartilage |
| 218(0.07) | 1(0.05) | NA | NA | |
| 26,372(8.64) | 289(8.94) | 1.04[0.87−1.26] | 1 | |
| 26,372(8.64) | 109(8.45) | 1.01[0.75−1.36] | 1 | C43−C44 Melanoma and other malignant neoplasms of skin |
| 26,372(8.64) | 171(9.26) | 1.06[0.84−1.35] | 1 | |
| 1,169(0.38) | 11(0.34) | 0.82[0.33−2.08] | 1 | |
| 1,169(0.38) | 3(0.23) | NA | NA | C45−C49 Malignant neoplasms of mesothelial and soft tissue |
| 1,169(0.38) | 8(0.43) | 1.13[0.40−3.18] | 1 | |
| 12,439(4.07) | 227(7.02) | 1.84[1.49−2.27] | 2.47E−12 | |
| 12,439(4.07) | 80(6.20) | 1.55[1.09−2.19] | 1.41E−02 | C50 Malignant neoplasm of breast |
| 12,439(4.07) | 139(7.53) | 2.05[1.57−2.69] | 4.49E−10 | |
| 4,047(1.33) | 50(1.55) | 1.20[0.79−1.83] | 1 | |
| 4,047(1.33) | 16(1.24) | 0.93[0.44−1.94] | 1 | C51−C58 Malignant neoplasms of female genital organs |
| 4,047(1.33) | 33(1.79) | 1.43[0.85−2.40] | 0.91 | |
| 10,557(3.46) | 179(5.53) | 1.77[1.39−2.25] | 3.64E−09 | |
| 10,557(3.46) | 58(4.50) | 1.52[1.01−2.30] | 0.14 | C60−C63 Malignant neoplasms of male genital organs |
| 10,557(3.46) | 113(6.12) | 1.87[1.38−2.53] | 4.13E−07 | |
| 4,351(1.43) | 77(2.38) | 1.75[1.24−2.46] | 1.01E−04 | |
| 4,351(1.43) | 27(2.09) | 1.58[0.89−2.80] | 0.84 | C64−C68 Malignant neoplasms of urinary tract |
| 4,351(1.43) | 49(2.65) | 1.92[1.25−2.94] | 2.68E−04 | |
| 1,031(0.34) | 14(0.43) | 1.30[0.59−2.84] | 1 | |
| 1,031(0.34) | 7(0.54) | 1.66[0.55−5.02] | 1 | C69−C72 Malignant neoplasms of eye, brain and other parts of central nervous system |
| 1,031(0.34) | 5(0.27) | 0.80[0.22−2.95] | 1 | |
| 773(0.25) | 18(0.56) | 2.23[1.11−4.49] | 0.08 | |
| 773(0.25) | 7(0.54) | 2.15[0.71−6.52] | 1 | C73−C75 Malignant neoplasms of thyroid and other endocrine glands |
| 773(0.25) | 10(0.54) | 2.19[0.86−5.56] | 0.2 | |
| 14,339(4.70) | 192(5.94) | 1.31[1.05−1.63] | 4.68E−03 | |
| 14,339(4.70) | 66(5.12) | 1.13[0.78−1.64] | 1 | C76−C79 Malignan neoplasms of ill−defined, secondary and unspecified sites |
| 14,339(4.70) | 120(6.50) | 1.43[1.09−1.90] | 2.95E−03 | |
| 5,030(1.65) | 108(3.34) | 2.11[1.58−2.82] | 3.45E−09 | |
| 5,030(1.65) | 39(3.02) | 1.95[1.21−3.15] | 4.70E−03 | C81−C96 Malignant neoplasms, stated or presumed to be primary, of lymphoid, haematopoietic and related tissue |
| 5,030(1.65) | 65(3.52) | 2.19[1.51−3.17] | 8.74E−07 | |

UK Biobank

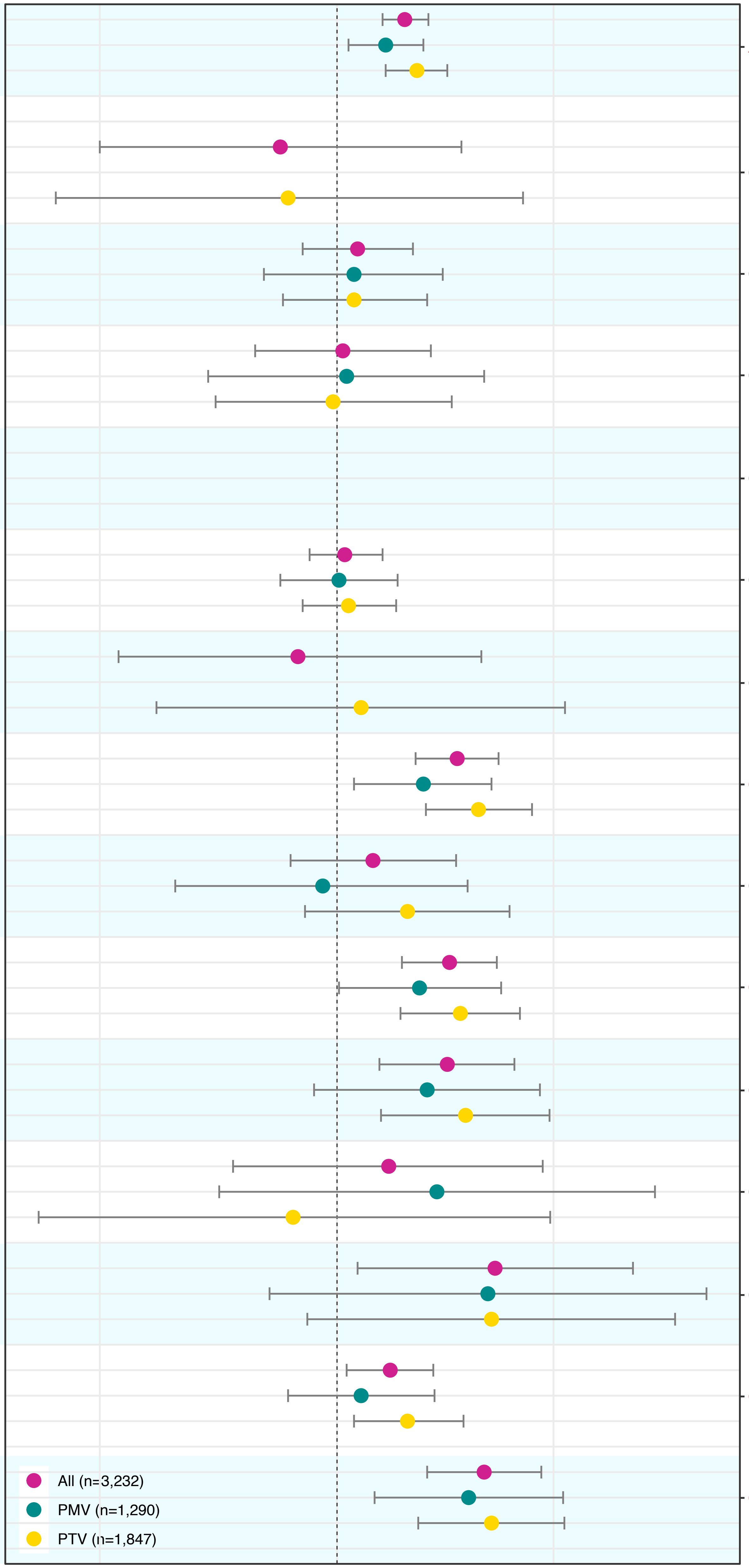
