## Supplemental Figure 4 for "Genomic ascertainment of *CHEK2*-related cancer predisposition"

| Controls(%) | Heterozygotes(%) | OR [95% CI] | Adjusted SAIGE p−value | Geisinger |
|---|---|---|---|---|
| 32(0.05) | 1(0.08) | NA | NA | |
| 32(0.05) | 0(0) | NA | NA | C60 Malignant neoplasm of penis |
| 32(0.05) | 1(0.08) | NA | NA | |
| 4222(7.12) | 135(11.09) | 1.62[1.27−2.07] | 1.04E−04 | |
| 4222(7.12) | 101(11.51) | 1.69[1.27−2.24] | 4.03E−04 | C61 Malignant neoplasm of prostate |
| 4222(7.12) | 34(10.14) | NA | NA | |
| 195(0.33) | 7(0.58) | 1.74[0.67−4.55] | 1 | |
| 195(0.33) | 6(0.68) | 2.06[0.73−5.83] | 0.43 | C62 Malignant neoplasm of testis |
| 195(0.33) | 1(0.298) | NA | NA | |
| 19(0.03) | 0(0) | NA | NA | |
| 19(0.03) | 0(0) | NA | NA | C63 Malignant neoplasm of other and unspecified male genital organs |
| 19(0.03) | 0(0) | NA | NA | |
| 1202(0.79) | 39(1.24) | 1.58[1.03−2.41] | 0.02 | |
| 1202(0.79) | 27(1.22) | NA | NA | C64 Malignant neoplasm of kidney, except renal pelviss |
| 1202(0.79) | 12(1.31) | 1.76[0.83−3.75] | 0.28 | |
| 120(0.08) | 4(0.13) | NA | NA | |
| 120(0.08) | 1(0.05) | NA | NA | C65 Malignant neoplasm of renal pelvis |
| 120(0.08) | 3(0.33) | NA | NA | |
| 90(0.06) | 3(0.10) | NA | NA | |
| 90(0.06) | 3(0.14) | NA | NA | C66 Malignant neoplasm of ureter |
| 90(0.06) | 0(0) | NA | NA | |
| 1477(0.96) | 46(1.46) | 1.50[1.01−2.23] | 0.1 | |
| 1477(0.96) | 27(1.22) | NA | NA | C67 Malignant neoplasm of bladder |
| 1477(0.96) | 19(2.08) | 2.34[1.25−4.38] | 3.51E−03 | |
| 193(0.13) | 6(0.19) | 1.47[0.51−4.27] | 1 | |
| 193(0.13) | 2(0.1) | NA | NA | C68 Malignant neoplasm of other and unspecified urinary organs |
| 193(0.13) | 4(0.44) | NA | NA | |
| 1370(0.90) | 57(1.81) | 2.04[1.48−2.82] | 1.82E−05 | |
| 1370(0.90) | 42(1.89) | 2.15[1.47−3.14] | 6.60E−05 | C73 Malignant neoplasm of thyroid gland |
| 1370(0.90) | 14(1.53) | NA | NA | |
| 80(0.05) | 3(0.10) | NA | NA | |
| 80(0.05) | 3(0.14) | NA | NA | C74 Malignant neoplasm of adrenal gland |
| 80(0.05) | 0(0) | NA | NA | |
| 371(0.24) | 6(0.19) | 0.79[0.30−2.10] | 1 | |
| 371(0.24) | 4(0.2) | NA | NA | C75 Malignant neoplasm of other endocrine glands and related structures |
| 371(0.24) | 2(0.22) | NA | NA | |
| 266(0.17) | 6(0.19) | 1.09[0.33−3.58] | 1 | |
| 266(0.17) | 3(0.14) | NA | NA | C81 Hodgkin lymphoma |
| 266(0.17) | 3(0.33) | NA | NA | |
| 397(0.26) | 15(0.47) | 1.80[0.84−3.84] | 0.5 | |
| 397(0.26) | 12(0.54) | NA | NA | C82 Follicular lymphoma |
| 397(0.26) | 3(0.33) | NA | NA | |
| 677(0.44) | 19(0.60) | 1.34[0.68−2.64] | 1 | |
| 677(0.44) | 11(0.50) | NA | NA | C83 Non−follicular lymphoma |
| 677(0.44) | 8(0.88) | 2.04[0.73−5.73] | 0.87 | |
| 160(0.10) | 4(0.13) | NA | NA | |
| 160(0.10) | 3(0.14) | NA | NA | C84 Periphera and cutaneous T−cell lymphomas |
| 160(0.10) | 1(0.11) | NA | NA | |
| 1216(0.80) | 35(1.11) | 1.38[0.84−2.28] | 1 | |
| 1216(0.80) | 24(1.08) | NA. | NA. | C85 Other specified and unspecified types of non−Hodgkin lymphoma |
| 1216(0.80) | 11(1.20) | 1.55[0.64−3.75] | 1 | |
| 22(0.01) | 0(0) | NA | NA | |
| 22(0.01) | 0(0) | NA | NA | C86 Other specified types of T/NK cell lymphoma |
| 22(0.01) | 0(0) | NA | NA | |
| 135(0.10) | 1(0.03) | NA | NA | |
| 135(0.10) | 1(0.05) | NA | NA | C88 Malignant immunoproliferative diseases |
| 135(0.10) | 0(0) | NA | NA | |
| 541(0.35) | 13(0.41) | 1.14[0.50−2.59] | 1 | |
| 541(0.35) | 6(0.27) | NA | NA | C90 Multiple myeloma and malignant plasma cell neoplasms |
| 541(0.35) | 7(0.77) | 2.23[0.74−6.74] | 0.36 | |
| 627(0.41) | 27(0.86) | 2.08[1.17−3.69] | 7.61E−03 | |
| 627(0.41) | 19(0.86) | NA | NA | C91 Lymphoid leukemia |
| 627(0.41) | 8(0.88) | 2.21[0.78−6.22] | 0.53 | |
| 338(0.22) | 12(0.38) | 1.72[0.74−4.02] | 1 | |
| 338(0.22) | 7(0.32) | NA | NA | C92 Myeloid leukemia |
| 338(0.22) | 5(0.54) | 2.53[0.69−9.33] | 0.57 | |
| 36(0.02) | 0(0) | NA | NA | |
| 36(0.02) | 0(0) | NA | NA | C93 Monocytic leukemia |
| 36(0.02) | 0(0) | NA | NA | |
| 35(0.02) | 0(0) | NA | NA | |
| 35(0.02) | 0(0) | NA | NA | C94 Other leukemia of specified cell type |
| 35(0.02) | 0(0) | NA | NA | |
| 147(0.09) | 6(0.19) | 1.95[0.58−6.52] | 0.3 | |
| 147(0.09) | 1(0.05) | NA | NA | C95 Leukemia of unspecified cell type |
| 147(0.09) | 4(0.44) | NA | NA | |
| 108(0.07) | 2(0.06) | NA | NA | |
| 108(0.07) | 1(0.05) | NA | NA | C96 Other and unspecified malignant_neoplasms_of_lymphoid,_hematopoietic_and_related_tissue |
| 108(0.07) | 1(0.11) | NA | NA | |

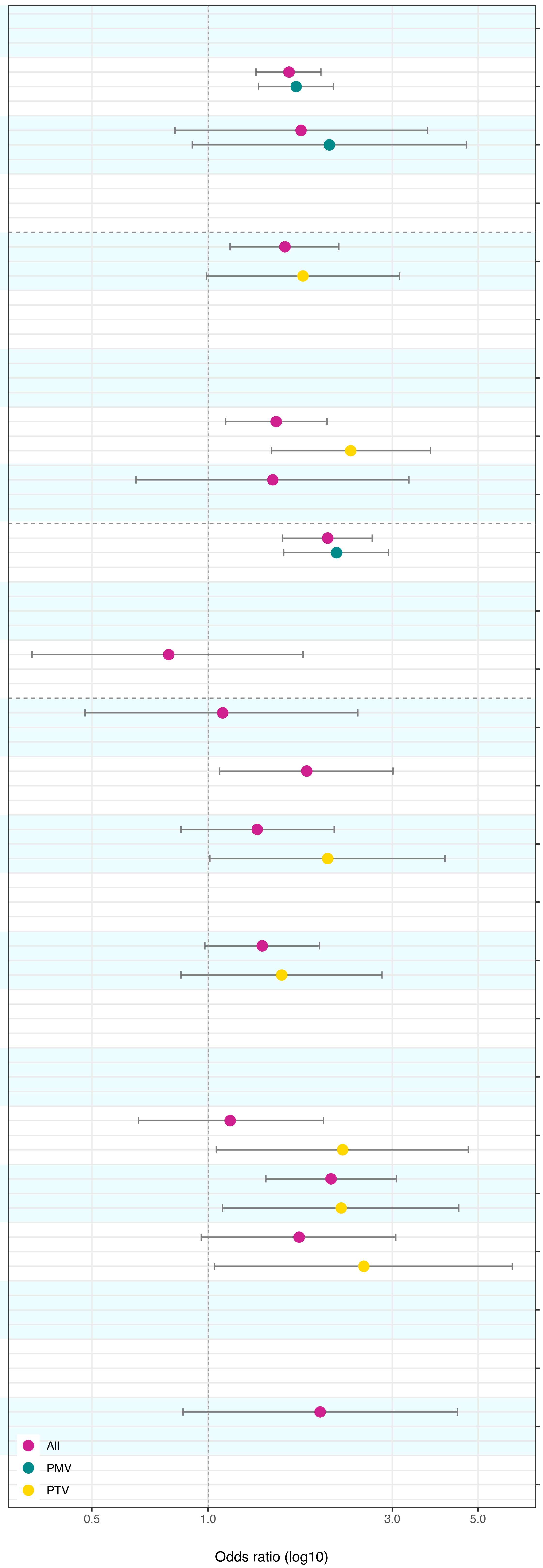
