## Supplemental Figure 5 for "Genomic ascertainment of *CHEK2*-related cancer predisposition"

| Disease | Controls(%) | Heterozygotes(%) | OR [95% CI] | Adjusted SAIGE p−value |
|---|---|---|---|---|
| C60 Malignant neoplasm of penis | 91(0.03) | 0(0) | NA | NA |
| | 91(0.03) | 0(0) | NA | NA |
| | 91(0.03) | 0(0) | NA | NA |
| C61 Malignant neoplasm of prostate | 9,978(3.27) | 170(5.27) | 1.78[1.48−2.16] | 6.68E−10 |
| | 9,978(3.27) | 55(4.26) | NA | NA |
| | 9,978(3.27) | 107(5.79) | 1.87[1.47−2.38] | 1.27E−07 |
| C62 Malignant neoplasm of testis | 517(0.17) | 10(0.31) | 1.82[0.89−3.72] | 0.12 |
| | 517(0.17) | 3(0.23) | NA | NA |
| | 517(0.17) | 7(0.38) | 2.16[0.92−5.09] | 0.15 |
| C63 Malignant neoplasm of other and unspecified male genital organs | 22(0.01) | 0(0) | NA | NA |
| | 22(0.01) | 0(0) | NA | NA |
| | 22(0.01) | 0(0) | NA | NA |
| C64 Malignant neoplasm of kidney, except renal pelvis | 1,635(0.54) | 31(0.96) | 1.84[1.22−2.77] | 2.49E−03 |
| | 1,635(0.54) | 11(0.85) | NA | NA |
| | 1,635(0.54) | 20(1.08) | 2.05[1.23−3.40] | 4.28E−03 |
| C65 Malignant neoplasm of renal pelvis | 142(0.05) | 2(0.06) | NA | NA |
| | 142(0.05) | 2(0.16) | NA | NA |
| | 142(0.05) | 0(0) | NA | NA |
| C66 Malignant neoplasm of ureter | 163(0.05) | 1(0.03) | NA | NA |
| | 163(0.05) | 1(0.08) | NA | NA |
| | 163(0.05) | 0(0) | NA | NA |
| C67 Malignant neoplasm of bladder | 2,707(0.89) | 45(1.40) | 1.64[1.17−2.31] | 2.66E−03 |
| | 2,707(0.89) | 15(1.16) | NA. | NA. |
| | 2,707(0.89) | 30(1.62) | 1.88[1.24−2.85] | 2.32E−03 |
| C68 Malignant neoplasms of other and unspecified unrinary organs | 86(0.06) | 3(0.09) | NA | NA |
| | 86(0.06) | 1(0.08) | NA | NA |
| | 86(0.06) | 2(0.11) | NA | NA |
| C76 Malignant neoplasm of other and ill−defined sites | 175(0.06) | 3(0.09) | NA | NA |
| | 175(0.06) | 1(0.08) | NA | NA |
| | 175(0.06) | 2(0.11) | NA | NA |
| C77 Secondary and unspecified malignant neoplasm of lymph nodes | 7,493(2.45) | 109(3.38) | 1.41[1.11−1.78] | 9.77E−04 |
| | 7,493(2.45) | 40(3.10) | NA | NA |
| | 7,493(2.45) | 66(3.57) | 1.50[1.11−2.03] | 4.23E−03 |
| C78 Secondary malignant neoplasm of respiratory and digestive organs | 7,557(2.48) | 94(2.92) | 1.21[0.94−1.55] | 0.21 |
| | 7,557(2.48) | 32(2.48) | NA | NA |
| | 7,557(2.48) | 59(3.19) | 1.32[0.96−1.81] | 0.16 |
| C79 Secondary malignant neoplasm of other sites | 6,072(2.00) | 84(2.61) | 1.34[1.03−1.76] | 0.01 |
| | 6,072(2.00) | 27(2.09) | NA | NA |
| | 6,072(1.99) | 56(3.03) | 1.56[1.13−2.17] | 4.40E−03 |
| C81 Hodgkin's disease | 414(0.14) | 8(0.25) | 1.83[0.68−4.94] | 0.02 |
| | 414(0.14) | 0(0) | NA | NA |
| | 414(0.14) | 8(0.43) | 3.19[1.22−8.36] | 8.40E−03 |
| C82 Follicular [nodular] non−Hodgkin's lymphoma | 593(0.19) | 10(0.31) | 1.61[0.66−3.90] | 1 |
| | 593(0.19) | 5(0.39) | 2.04[0.61−6.84] | 1 |
| | 593(0.19) | 5(0.27) | 1.39[0.42−4.67] | 1 |
| C83 Diffuse non−Hodgkin's lymphoma | 1,362(0.45) | 26(0.81) | 1.84[1.06−3.20] | 0.03 |
| | 1,362(0.45) | 7(0.54) | 1.27[0.46−3.55] | 1 |
| | 1,362(0.45) | 18(0.97) | 2.19[1.15−4.17] | 0.01 |
| C84 Peripheral and cutaneous T−cell lymphomas | 216(0.07) | 6(0.19) | 2.68[0.85−8.45] | 0.24 |
| | 216(0.07) | 5(0.39) | 5.76[1.70−19.50] | 0.02 |
| | 216(0.07) | 1(0.05) | NA | NA |
| C85 Other and unspecified types of non−Hodgkin's lymphoma | 1,533(0.50) | 29(0.90) | 1.83[1.08−3.08] | 0.02 |
| | 1,533(0.50) | 8(0.62) | 1.28[0.49−3.34] | 0.94 |
| | 1,533(0.50) | 20(1.08) | 2.18[1.18−4.00] | 0.02 |
| C86 Other specified types of T/NK−cell lymphoma | 42(0.01) | 4(0.12) | NA | NA |
| | 42(0.01) | 3(0.23) | NA | NA |
| | 42(0.01) | 1(0.05) | NA | NA |
| C88 Malignant immunoproliferative diseases | 180(0.06) | 4(0.12) | NA | NA |
| | 180(0.06) | 1(0.08) | NA | NA |
| | 180(0.06) | 3(0.16) | NA | NA |
| C90 Multiple myeloma and malignant plasma cell neoplasms | 933(0.31) | 18(0.56) | 1.86[0.96−3.61] | 0.09 |
| | 933(0.31) | 7(0.54) | 1.87[0.67−5.20] | 1 |
| | 933(0.31) | 10(0.54) | 1.77[0.75−4.19] | 0.29 |
| C91 Lymphoid leukaemia | 921(0.30) | 21(0.65) | 2.21[1.19−4.08] | 0.01 |
| | 921(0.30) | 9(0.70) | 2.44[0.99−6.05] | 0.14 |
| | 921(0.30) | 12(0.65) | 2.16[0.99−4.74] | 0.08 |
| C92 Myeloid leukaemia | 650(0.21) | 20(0.62) | 2.97[1.58−5.58] | 2.17E−04 |
| | 650(0.21) | 7(0.54) | 2.66[0.95−7.41] | 0.13 |
| | 650(0.21) | 13(0.70) | 3.32[1.56−7.08] | 1.22E−04 |
| C93 Monocytic leukaemia | 82(0.03) | 2(0.06) | NA | NA |
| | 82(0.03) | 2(0.16) | NA | NA |
| | 82(0.03) | 0(0) | NA | NA |
| C94 Other leukaemias of specified cell type | 46(0.02) | 3(0.09) | NA | NA |
| | 46(0.02) | 2(0.16) | NA | NA |
| | 46(0.02) | 1(0.05) | NA | NA |
| C95 Leukaemia of unspecified cell type | 103(0.03) | 1(0.03) | NA | NA |
| | 103(0.03) | 0(0) | NA | NA |
| | 103(0.03) | 1(0.05) | NA | NA |
| C96 Other and unspecified malignant neoplasms of lymphoid, haematopoietic and related tissue | 50(0.02) | 5(0.16) | 9.47[2.58−34.80] | 1.91E−03 |
| | 50(0.02) | 3(0.23) | NA | NA |
| | 50(0.02) | 2(0.11) | NA | NA |

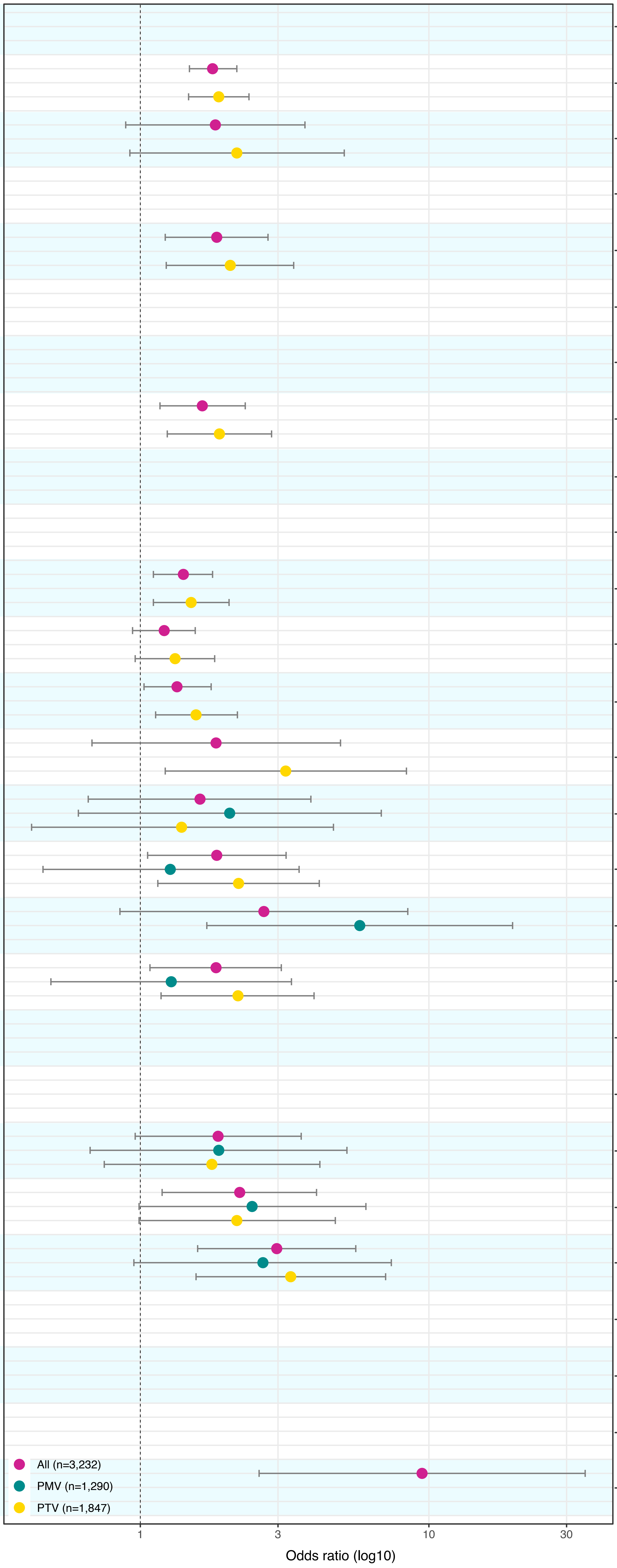
